# Sleep Regularity Index as a Novel Indicator of Sleep Disturbance in Stroke Survivors: A Secondary Data Analysis

**DOI:** 10.1101/2024.11.19.24317536

**Authors:** Katrijn B. Schruers, Matthew Weightman, Anna á V. Guttesen, Barbara Robinson, Heidi Johansen-Berg, Melanie K. Fleming

**Affiliations:** Wellcome Centre for Integrative Neuroimaging (WIN), FMRIB, Nuffield Department of Clinical Neurosciences, University of Oxford; Faculty of Psychology and Neurosciences, Maastricht University, Maastricht, The Netherlands

**Author notes:** Corresponding author: Melanie Fleming, Wellcome Centre for Integrative Neuroimaging John Radcliffe Hospital, Oxford OX3 9DU United Kingdom.

## Abstract

**Background:** Sleep disturbance is common but often overlooked following stroke. Recent studies have highlighted the importance of sleep regularity in overall health, however, there is little information about sleep regularity after stroke. This study aimed to test for differences in the sleep regularity index (SRI), derived from actigraphy data, between stroke survivors and healthy controls. Secondary objectives included testing for correlations between SRI and other actigraphy derived sleep metrics in both groups, and the association between SRI and depression, disability, quality of life, and chronicity in the stroke group.

**Methods:** Actigraphy data were obtained from an existing dataset (approx. 1 week of data per participant). SRI calculation followed established procedures for 162 community-dwelling stroke survivors (mean age 61±14 years, 5±5 years post-stroke, 89 males) and 60 healthy controls (mean age 57±17 years, 32 males). The primary outcome measure was SRI (score 0-100; higher scores indicating greater sleep regularity). Other sleep metrics included total sleep time, fragmentation, sleep efficiency, wake after sleep onset and self-reported sleep.

**Results:** The SRI was significantly lower for stroke survivors compared to healthy controls (*p*=0.001). Higher SRI correlated with longer total sleep time (*p*=0.003) and better self-reported sleep (sleep condition indicator; *p*=0.001) for the stroke group, but not for any other sleep metrics, nor for controls. For the stroke group, lower SRI was associated with worse depression (*p*=0.006), and quality of life (*p*=0.001), but there were no associations with post-stroke disability (*p*=0.886) nor time since stroke (*p*=0.646).

**Conclusion:** This study highlights potential disrupted sleep regularity post-stroke. Future research should explore interventions targeting sleep regularity to improve sleep quality and overall outcomes in this population.

## Introduction

Sleep-related impairments are a prevalent, yet often overlooked, complication following stroke^1^. Diminished subjective (self-reported) and objective (polysomnography) sleep quality has been commonly reported in comparison with control groups^2,3^. This is important as disruptions in sleep are thought to exacerbate coexisting symptoms, such as cognitive deficits, fatigue, and depression. Moreover, these sleep disturbances may impede rehabilitation trajectories and hinder an individual’s capacity to reintegrate into society^4,5^.

To investigate sleep in stroke survivors, multiple approaches can be taken. Polysomnography (PSG), the gold standard for sleep monitoring, records physiological changes that occur during sleep. However, PSG is typically conducted in sleep clinics utilising specialised equipment and trained personnel, rendering it costly and difficult to access^6^. In comparison, actigraphy, which is used to infer sleep based on movement recorded from a sensor on a limb, has demonstrated good sensitivity and accuracy similar to PSG^7,8^. High sensitivity refers to the ability of actigraphy to accurately detect periods of sleep, while its lower specificity indicates a reduced capacity to correctly identify periods of wakefulness. Despite lower specificity than PSG, actigraphy benefits from its capability to be worn continuously in the home environment over multiple days/nights, allowing for the investigation of longer-term sleep patterns^2,6,9^. In addition to assessing sleep, by leveraging around-the-clock data, actigraphy can also be used to investigate circadian rhythm alterations. This is important as both sleep disturbances and circadian rhythm dysfunctions can be risk factors for, and consequences of, stroke^10^. The circadian rhythm, responsible for controlling the sleep-wake cycle and regulating physiological processes, is vital for overall health^11^. Laboratory studies have demonstrated that misalignment between the timing of the sleep-wake cycle and endogenous circadian rhythmicity can impair attention, neurobehavioral performance, mood, and cognition^12^. These findings highlight the potential impact of sleep regularity on various aspects of health and well-being of relevance to stroke recovery.

In 2017, Phillips and colleagues^13^ introduced a new metric to measure sleep regularity with actigraphy, named the Sleep Regularity Index (SRI). The SRI is defined as the probability of an individual being in the same state (asleep or awake) at any two time points 24 hours apart.

Unlike other actigraphy measures, the SRI does not rely on a single main sleep period, making it suitable for individuals with multiple sleep periods throughout the day^13^. Lunsford-Avery and colleagues^14^ used the SRI in older adults, finding that less regular sleep was linked to increased cardiometabolic risk, delayed sleep timing, increased daytime sleep and sleepiness, and reduced light exposure, independently of sleep duration. Additionally, they established a link between sleep irregularity and worse depression symptoms in older adults. Furthermore, it has been established that sleep regularity, as measured by the SRI, is more strongly associated with all-cause mortality risk than sleep duration^15^. Taken together, these results underscore the importance of circadian rhythm continuity to overall health: however, the SRI has not been investigated in stroke survivors. Exploring the impact of stroke on sleep regularity and its association with other post-stroke effects could enhance our understanding and management of sleep-related issues in stroke patients. We therefore aimed to investigate the SRI in stroke survivors to better understand the differences in comparison to healthy controls, the relationship to self-reported sleep and actigraphy sleep metrics, as well as explore the potential correlations with disability, depression, and quality of life. Our primary hypothesis was that there would be a significant difference in SRI between stroke survivors and healthy controls, with a lower SRI (i.e., less regular sleep) for stroke survivors. We also hypothesised that SRI would correlate significantly with self-reported sleep and actigraphy sleep metrics (sleep fragmentation, sleep efficiency, Wake After Sleep Onset) but not with the Total Sleep Time.

## Methods

### Sample

We utilised previously acquired datasets at the Wellcome Centre for Integrative Neuroimaging (WIN), University of Oxford. Stroke patients and healthy controls (aged >18 years) were recruited for several studies spanning from 2017 to April 2024^2,16–18^. All participants included in the analysis provided informed consent at the time of their initial involvement. All five studies received ethical approval from either the National Research Ethics Service or by the University of Oxford Central University Research Ethics Committee (11/H0605/12, R40803, 22/EM/0080, R85306, 22/LO/0353). We only included participants living in the community, as sleep can be significantly affected by the hospital environment^19^. An overview of the studies and their respective inclusion/exclusion criteria are in supplementary Table S1.

As SRI has not previously been investigated in this population, there are no existing studies to guide sample size estimations. Thus, a sensitivity power analysis was performed (calculation of effect size given alpha, power, and available sample size). The data available at the start of the analysis included 184 stroke patients and 70 controls. Therefore, to perform an analysis of covariance (ANCOVA) for the primary outcome to detect the difference in SRI for stroke survivors compared to controls with covariates age and sex, the smallest effect size detectable was f=0.17 (G*Power v.3.1; alpha=0.05, power=80%, n=254 analysis of covariance).

### Measures Actigraphy

Across all studies we used a Motionwatch 8 device^20^, with participants instructed to wear it for approximately one week. If able, participants were directed to press the event marker both when initiating sleep in the evening and upon waking in the morning. They were also asked to complete a 7-night sleep diary, recording their attempted sleep and final wake times. Stroke survivors were instructed to wear the monitor on their less-affected wrist, controls and non-motor impaired participants were instructed to wear the monitor on their non-dominant wrist. In the current study, the single axis algorithm and peak detection recording mode of the MotionWatch 8 is used, as recommended for sleep recording with this device.

Actigraphy data were automatically classified as sleep or wake for each 30 second interval by the MotionWare programme. The start and end of the sleep period was based on a combination of information from the sleep diary, activity, ambient light, and the utilisation of the event marker. These data were used to calculate the following sleep metrics: Total Sleep Time (TST), sleep fragmentation index, Wake After Sleep Onset (WASO), and sleep efficiency (a full breakdown of how these metrics were calculated can be found in the supplementary materials).

### Calculation of SRI

The Sleep Regularity Index (SRI) used epoch data from actigraphy to calculate the percentage probability of an individual being in the same state (asleep vs. awake) at any two time-points 24 h apart, averaged across the study ^13^. The practical scale of the index ranges from 0 (completely random sleep/wake patterns) to 100 (perfectly consistent sleep/wake patterns) ^21^ and can then be used to classify individuals as Regular (top quintile) or Irregular (bottom quintile) sleepers ^13^. The current computation utilizes the 30-second epochs extracted from MotionWare and is in accordance with the calculations from Lunsford Avery et al ^14^. Considering N days of recording segmented into M daily epochs, *S*_*i*,*j*_=1 indicates the participant’s sleep status on day *i* during epoch *j*, and *S*_*i*,*j*_=0 indicates wakefulness. The SRI is then computed using the following equation (1), where *δ*(*S*_*i*,*j*_, *S*_*i*+1,*j*_) = 1 if *S*_*i*,*j*_ = *S*_*i*+1,*j*_ and 0 otherwise.

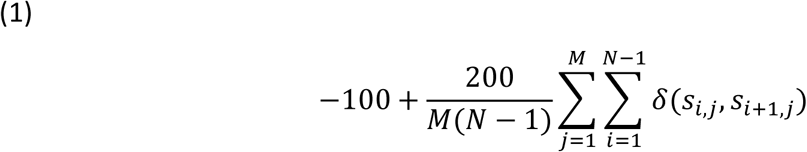

To guarantee a reliable calculation of the SRI, several criteria have been reported^21^ (see supplementary materials). These criteria were visually inspected per participant. Where these criteria were not satisfied, the recording was ruled invalid and excluded from analysis. 36 participants (26 patients/10 controls) were excluded based on these criteria, resulting in a final sample size of 222 (162 patients/60 controls).

Actigraphy demonstrates greater sensitivity compared to specificity, indicating proficiency in identifying periods of sleep, but relative ineffectiveness in accurately detecting wakefulness^22^. The SRI is unique in that it considers both daytime and night-time activity, necessitating a resolution to this inaccuracy while maintaining the distinctive capability to detect periods of sleep during the day, e.g. naps. To address this issue and enhance the reliability of recordings for SRI calculations, specialised algorithms can be implemented; the current study used the UCSD algorithm (with a scaling factor of 0.10) and applied Webster’s rescoring rules. To ensure transparency and accurate comparison between research studies, our procedure and refinements are detailed in supplementary materials (S2).

### Questionnaires

Self-reported sleep and depression were assessed with the Sleep Condition Indicator (SCI; range 0-32 (scores ≤ 16 indicative of probable insomnia disorder)^23^ and the Patient Health Questionnaire (PHQ-8; range 0-24, higher score indicates more severe depressive symptoms)^24^, respectively. Post-stroke disability was assessed with the modified Rankin Scale (mRS; score range 0= “no symptoms at all” to 6=“death”)^25^. Finally, quality of life was assessed with short-form Stroke Impact Scale (SF-SIS; max score 100, higher score indicates better quality of life)^26^.

### Statistical analysis

Analyses were conducted with Python (v.3.10.12), RStudio (R v.4.3.2) and MotionWare (v.1.3.33). Plots were created with GraphPad Prism (v.10.2.3). To compare age between groups, a Welch Two Sample t-test was performed, as it accommodates unequal variances and different sample sizes. To test the relationship between pairs of categorical variables (e.g. sex and group) a chi-square test was performed. To test the primary hypothesis that stroke survivors have a lower SRI than controls, an analysis of covariance (ANCOVA) was planned, with age and sex as covariates. However, the assumption of non-interaction in an ANCOVA model was violated. Therefore, a linear regression model was used instead. Testing of assumptions and further details can be found in supplementary materials (S3). To test whether the SRI relates to standard questionnaire and actigraphy-derived sleep metrics Pearson’s correlations were used (adjusted for age and sex) with Bonferroni correction for the multiple comparisons of the sleep metrics (α = 0.01; analysis details in supplementary S3). To ascertain if the strength of these correlations differs between groups, a Fisher’s R to Z-test was performed. Interpretation of Pearson’s correlations coefficients are reported according to the following guidelines: *r* < 0.4, weak; 0.4 ≤ *r* ≤ 0.7, moderate; and *r* > 0.7, strong^27^, all confidence Intervals (CIs) are reported at the 95% level and p-values were computed using a Wald t-distribution approximation.

## Results

### Demographics and descriptive statistics SRI

The final dataset included 162 stroke survivors (mean (±sd) 61 (± 14) years of age, 5 (± 5) years post-stroke, 89 males) and 60 healthy controls (57 (±17) years of age, 32 males; table 1). There was no significant difference in age (t(89.06) = 1.51, *p* = 0.135) or sex distribution (χ^2^ = 3.78e- 03, *p* = 0.951) between the groups.

**Table 1.**
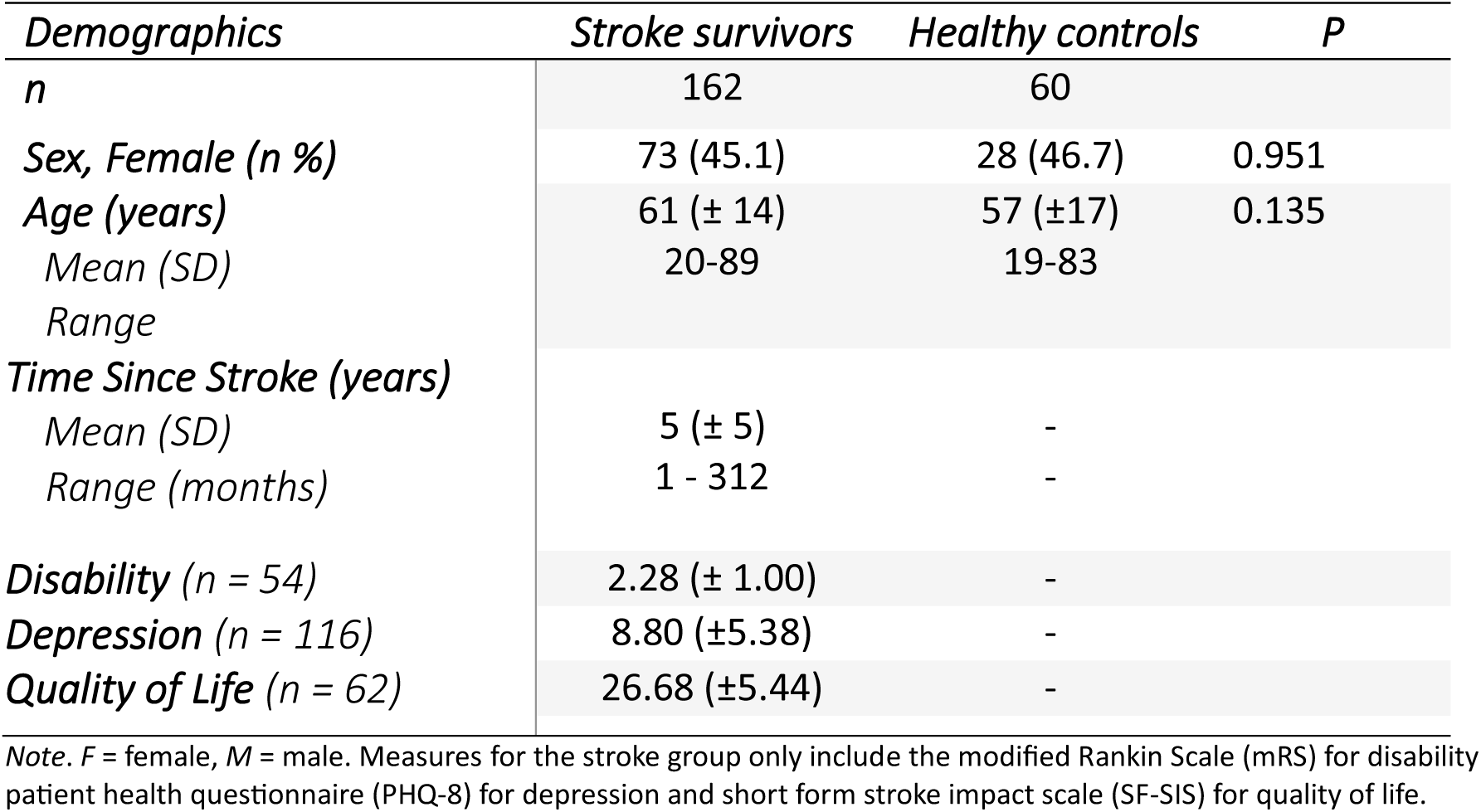

| *Demographics* | *Stroke survivors* | *Healthy controls* | *P* |
| --- | --- | --- | --- |
| *n* | 162 | 60 |  |
| *Sex, Female (n %)* | 73 (45.1) | 28 (46.7) | 0.951 |
| *Age (years)* | 61 (± 14) | 57 (±17) | 0.135 |
| *Mean (SD)* | 20-89 | 19-83 |  |
| *Range* |  |  |  |
| *Time Since Stroke (years)* |  |  |  |
| *Mean (SD)* | 5 (± 5) | - | |
| *Range (months)* | 1 - 312 | - |  |
| *Disability (n = 54)* | 2.28 (± 1.00) | - | |
| *Depression (n = 116)* | 8.80 (±5.38) | - | |
| *Quality of Life (n = 62)* | 26.68 (±5.44) | - | |

*Note.* F = female, M = male. Measures for the stroke group only include the modified Rankin Scale (mRS) for disability patient health questionnaire (PHQ-8) for depression and short form stroke impact scale (SF-SIS) for quality of life.

The mean (±sd) SRI for the stroke cohort was 41.21 (±12.58) and values ranged from 16.03 to 74.00. For the healthy controls, the mean (±sd) was 47.80 (±11.74) and the values ranged from 21.72 to 72.94 (figure 1).

**Figure 1.**
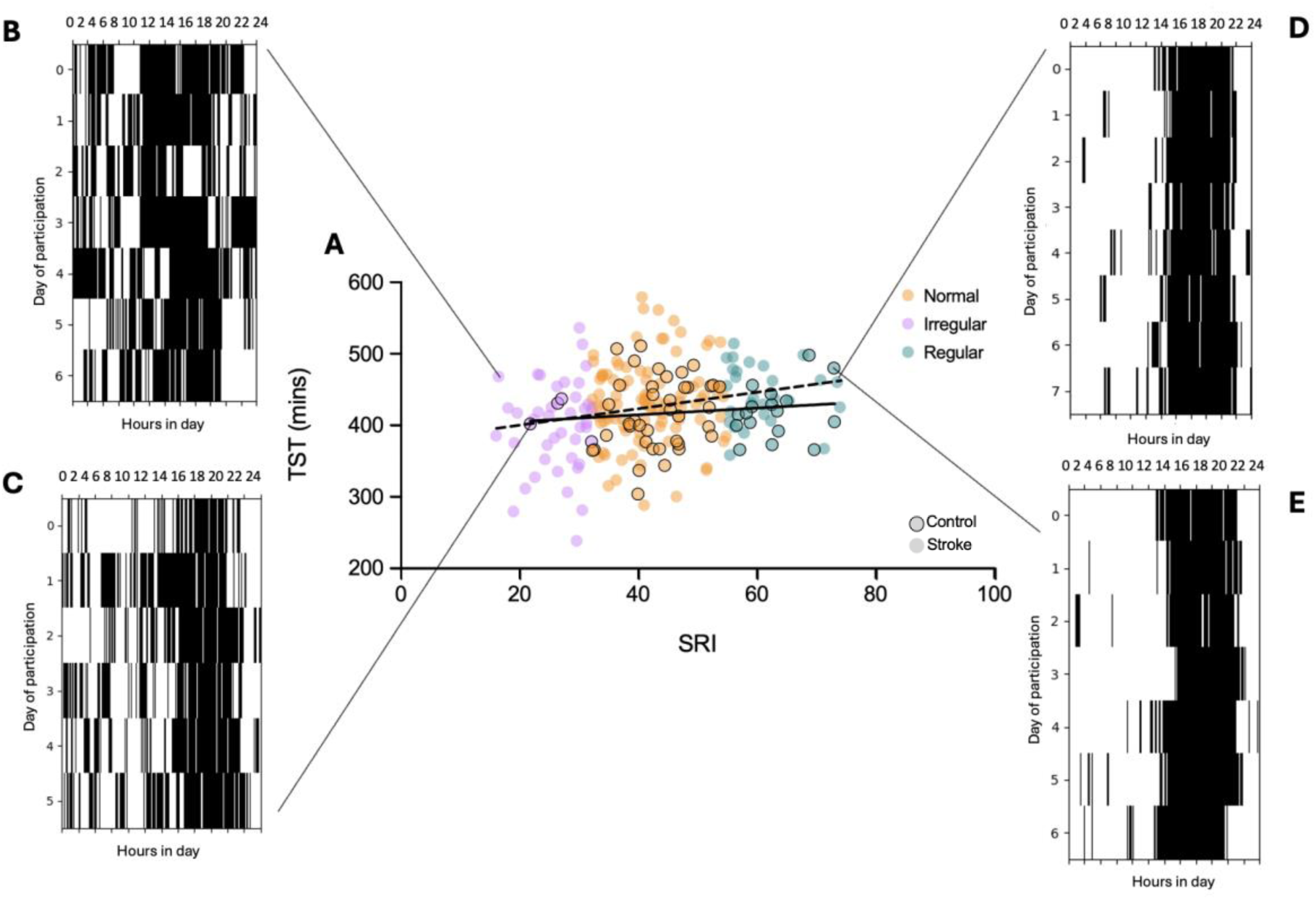
**A:** Relationship between sleep regularity (SRI) and total sleep time (TST) for the stroke cohort (dashed line) and healthy controls (solid line). Plots that show the sleep/wake cycle according to the SRI calculation for four participants, two classified as irregular sleepers (B=stroke, C=control), with SRI values in the first quintile, and two regular sleepers (D=stroke, E=control), with SRI values in the fifth quintile. The colours reflect ‘irregular’ (pink), ‘normal’ (orange) and ‘regular’ (teal) sleepers. The figure displays results from the current analysis, though the design is based on similar figures created by other studies^13,14^.

### Difference in sleep regularity between stroke survivors and healthy controls

SRI values are shown in Figure 2. To test for differences between groups, a linear regression model employing Ordinary Least Squares (OLS) estimation was utilised to predict SRI based on the factors Group (0 = Control, 1= Stroke), Age, and Sex. The linear regression model accounted for a significant but weak proportion of the variance in SRI (R^2^ = 0.12, F(3, 218) = 10.00, *p* < 0.001, adj. R^2^ = 0.11). Within this model, the effect of Group was significant and negative (β = -6.07, 95% CI [-9.65, -2.48], t(218) = -3.33, *p* = 0.001; Std. β = -0.48, 95% CI [-0.76, -0.20]), indicating that stroke survivors had a lower SRI, and thus less regular sleep compared to controls, as hypothesised. The effect of Age was significant and negative (β = - 0.12, 95% CI [-0.23, -0.01], t(218) = -2.16, *p* = 0.032; Std. β = -0.14, 95% CI [-0.27, -0.01]) demonstrating that increased age is associated with less regular sleep. The effect of Sex was significant and positive (β = 5.07, 95% CI [1.86, 8.29], t(218) = 3.11, *p* = 0.002; Std. β = 0.20, 95% CI [0.07, 0.33]), demonstrating that, overall, women have a higher SRI (more regular sleep) compared to men. Permutation testing (i.e., randomly subsampling the stroke group to account for the unequal sample sizes) revealed similar results (supplementary S4).

**Figure 2.**
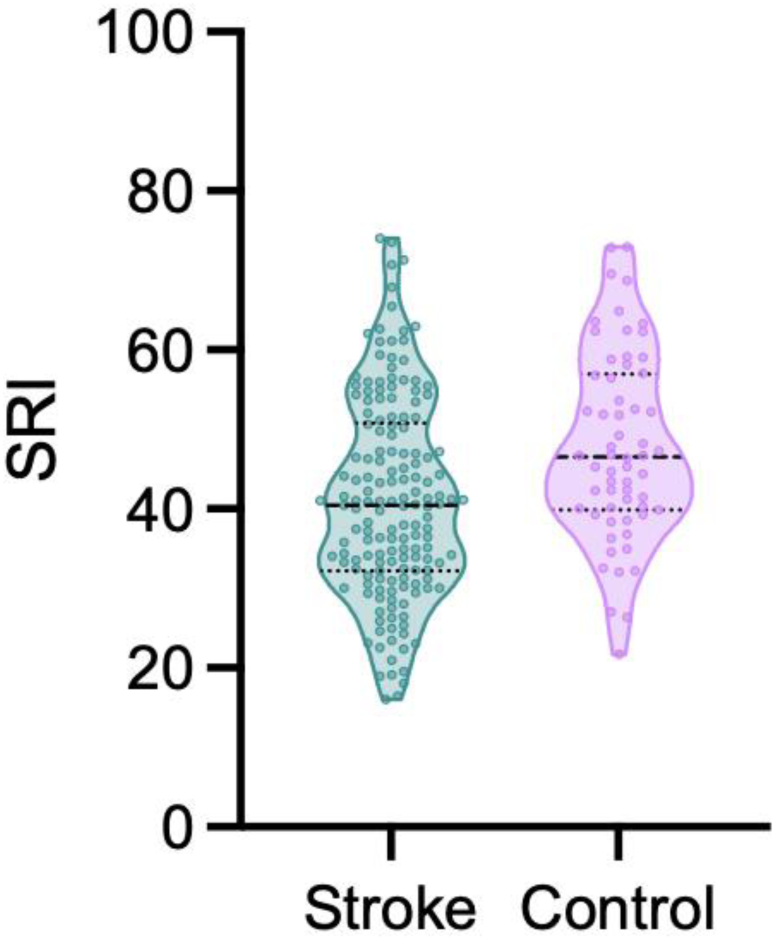
Violin plot showing the distribution of SRI scores in the stroke survivors and healthy controls. Higher SRI is indicative of more regular sleep. There was a significant effect of group, adjusting for age and sex (*p* = 0.001).

As the current research incorporates data from five distinct stroke studies, each with individualised objectives, there is an inherent sampling bias. In particular, several studies^16,17^ recruited stroke survivors based on their subjective experience of sleep issues and interest in receiving sleep treatment. We found a significant correlation between SCI and the SRI (with stroke and control combined; r = 0.25, *p* < 0.001; Figure 3), indicating the SRI is higher for participants with better self-reported sleep. We therefore repeated the linear regression with SCI as a predictor, to control for any differences between groups. The effect of Group is negative but no longer significant (β = -2.22, 95% CI [-6.29, 1.85], t(217) = -1.07, *p* = 0.284; standardized β = -0.18, 95% CI [-0.50, 0.15]).

**Figure 3.**
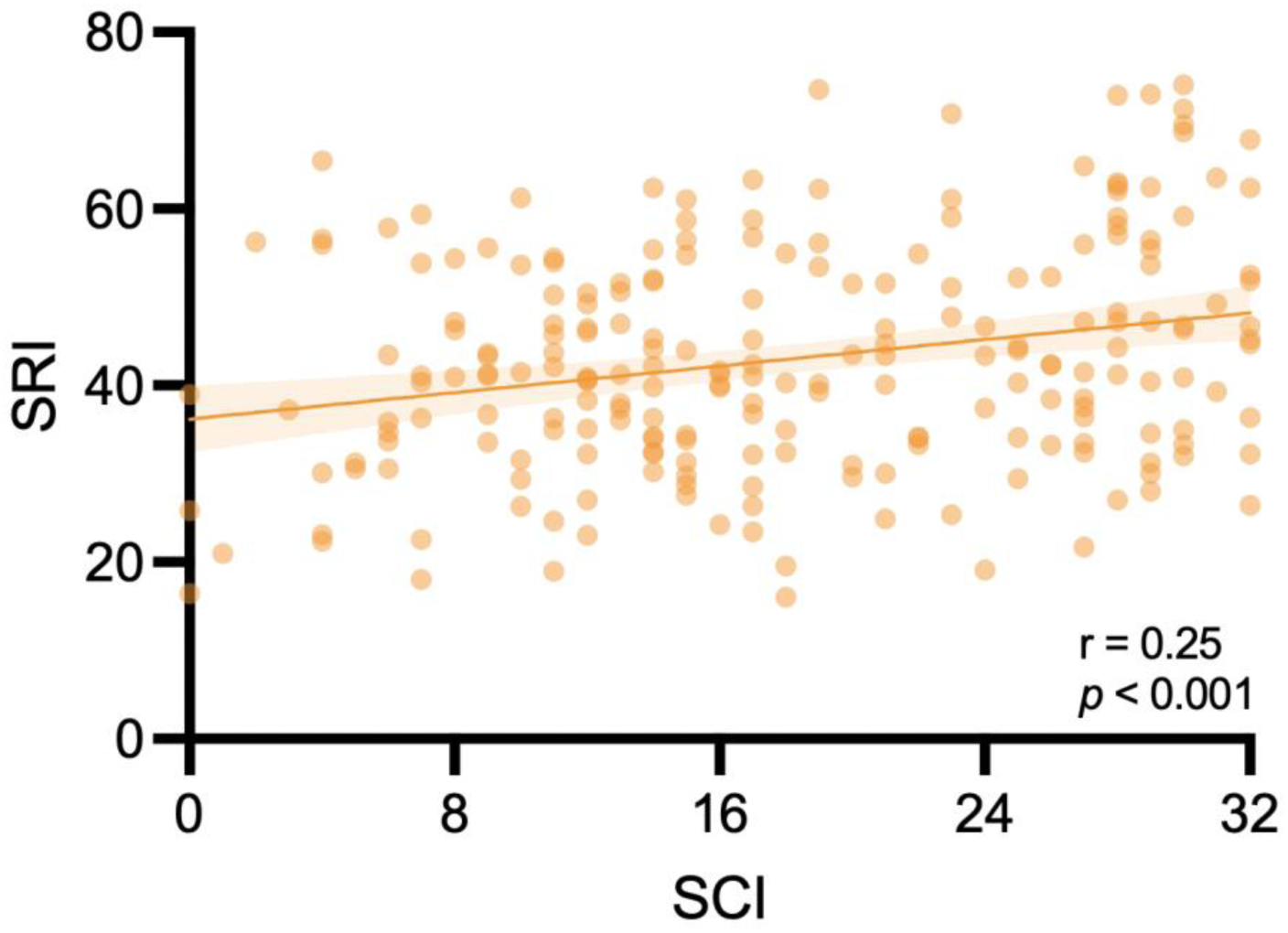
A scatterplot of the correlation between SCI and SRI for the whole dataset (including both stroke survivors and healthy controls), with better self-reported sleep associated with more regular sleep: r=0.25, *p* < 0.001.

**Figure 4.**
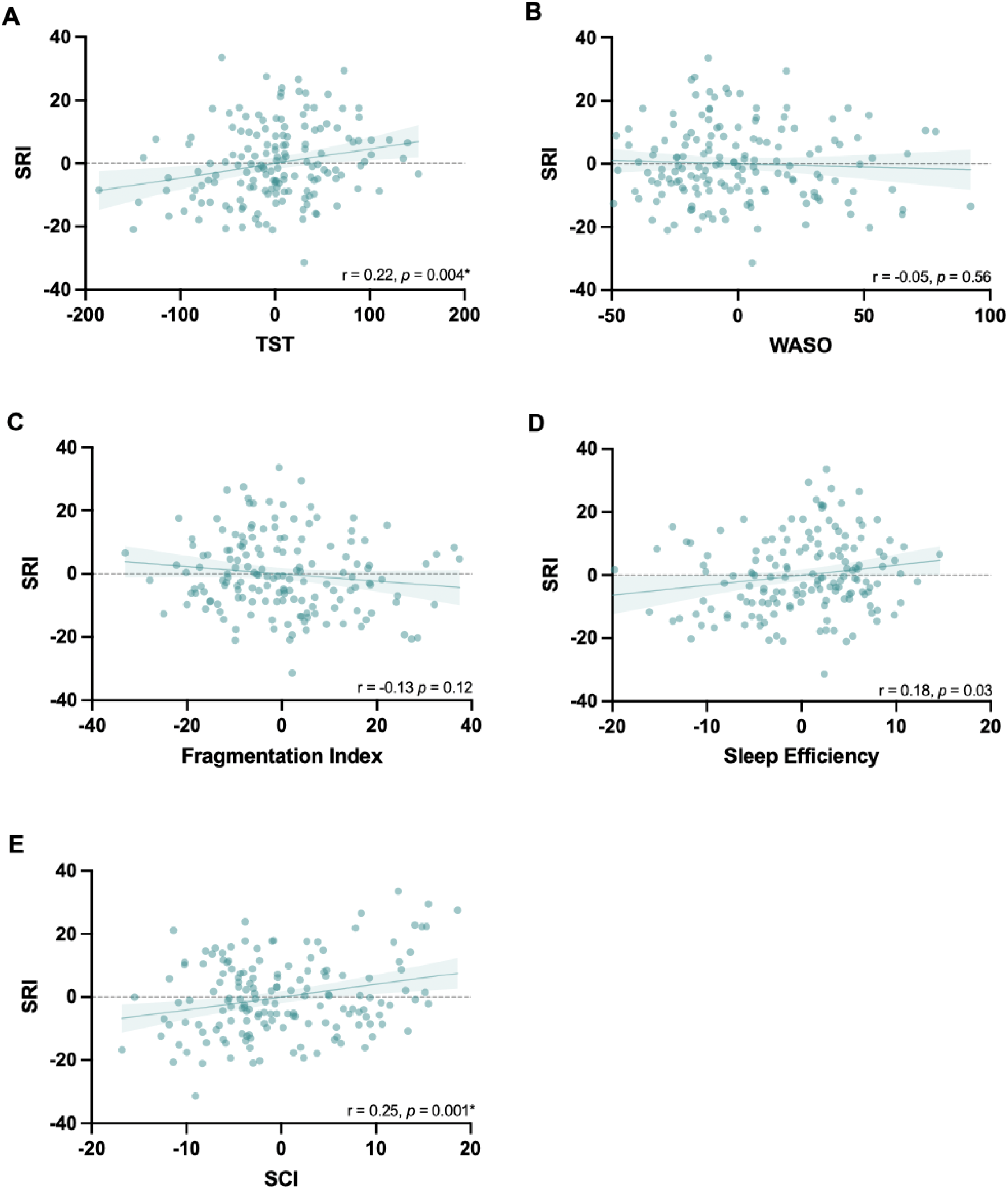
Plotted residuals accounted for age and sex to demonstrate partial correlations for the stroke cohort between the sleep regularity index (SRI) and **A.** total sleep time (TST), **B.** wake after sleep onset (WASO), **C.** fragmentation index, **D.** sleep efficiency, **E.** sleep condition indicator (SCI), α = 0.01 (Bonferroni corrected)

### Investigating correlations between SRI and other sleep metrics Stroke survivors

Counter to our hypothesis, there was a weak but statistically significant correlation between SRI and Total Sleep Time (r = 0.22, *p* = 0.004), indicating that when stroke survivors have more regular sleep, they have longer overall nighttime sleep. There was a weak but significant correlation between SRI and Sleep Condition Indicator score (r = 0.25, *p* = 0.001), suggesting that better self-reported sleep after stroke is associated with more regular sleep-wake patterns. However, there were no significant correlations between SRI and Wake After Sleep Onset (r = -0.05, *p* = 0.56), Fragmentation Index (r = -0.13, *p* = 0.12) nor Sleep Efficiency (r = 0.18, *p* = 0.03; *ns with Bonferroni correction*).

### Healthy controls

There were no correlations between SRI and TST (r = 0.07, *p* = 0.616), Fragmentation Index (r = 0.04, *p* = 0.792), Sleep efficiency (r = -0.06, *p* = 0.659), wake after sleep onset (r = 0.01, *p* = 0.943), nor the sleep condition indicator (r = 0.28, *p* = 0.040; *ns with Bonferroni correction*) in the control group (Figure 5). Comparisons of the strength of correlations between the groups revealed no statistically significant differences (supplementary S4).

**Figure 5.**
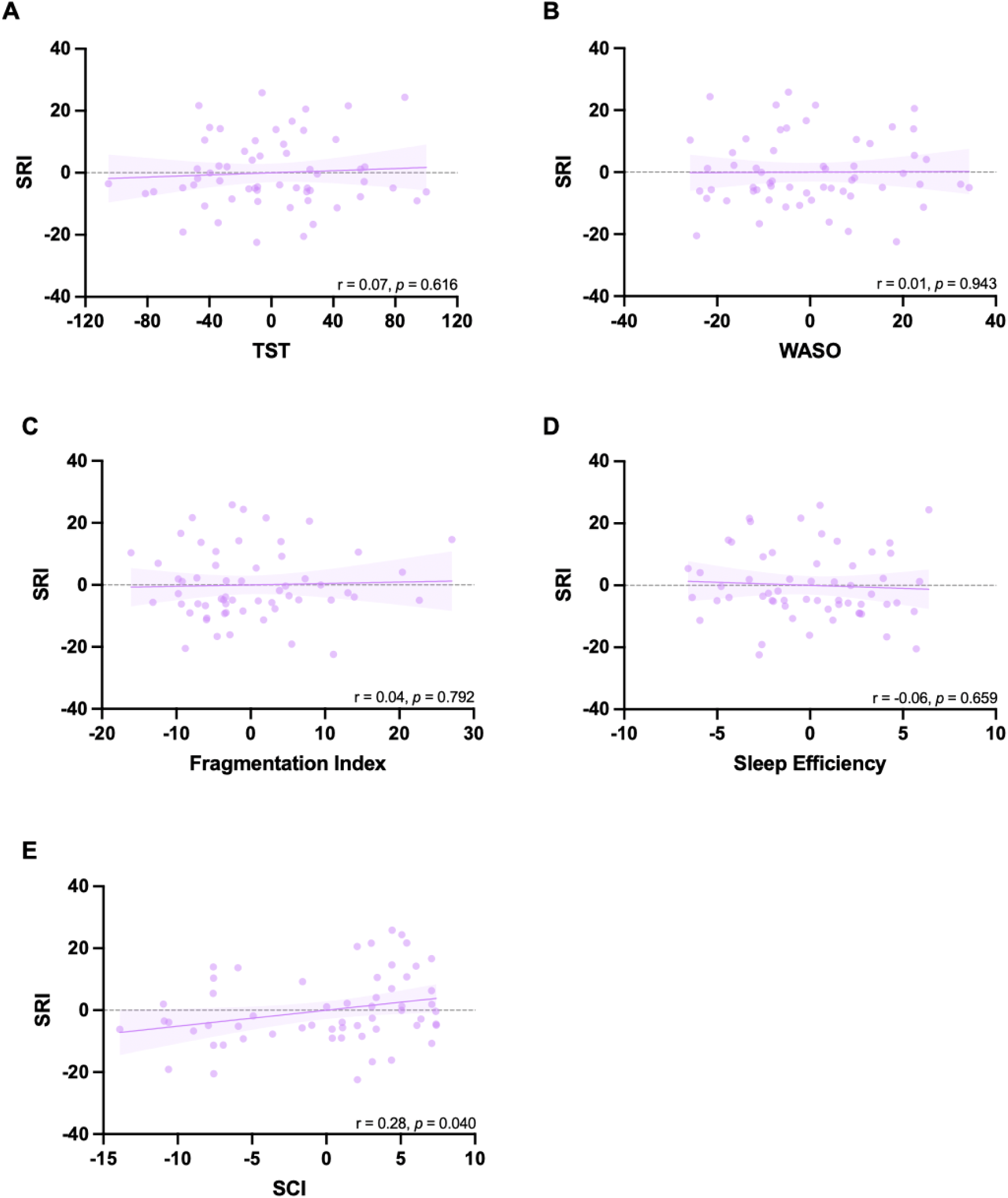
Plotted residuals accounted for age and sex to demonstrate partial correlations for the healthy control cohort between the sleep regularity index (SRI) and **A.** and total sleep time (TST), **B.** wake after sleep onset (WASO), **C.** fragmentation index, **D.** sleep efficiency, **E.** sleep condition indicator (SCI), α = 0.01 (Bonferroni correction for multiple comparisons).

### Stroke group analyses

To test the secondary hypothesis that stroke survivors with less regular sleep would have higher (worse) depression scores, a Pearson’s correlation (adjusted for age and sex) was performed on available data (n=113). There was a significant correlation between the SRI and PHQ-8 (r = -0.26, *p* = 0.006; Figure 6A) indicating that less regular sleep is associated with worse depression scores.

**Figure 6.**
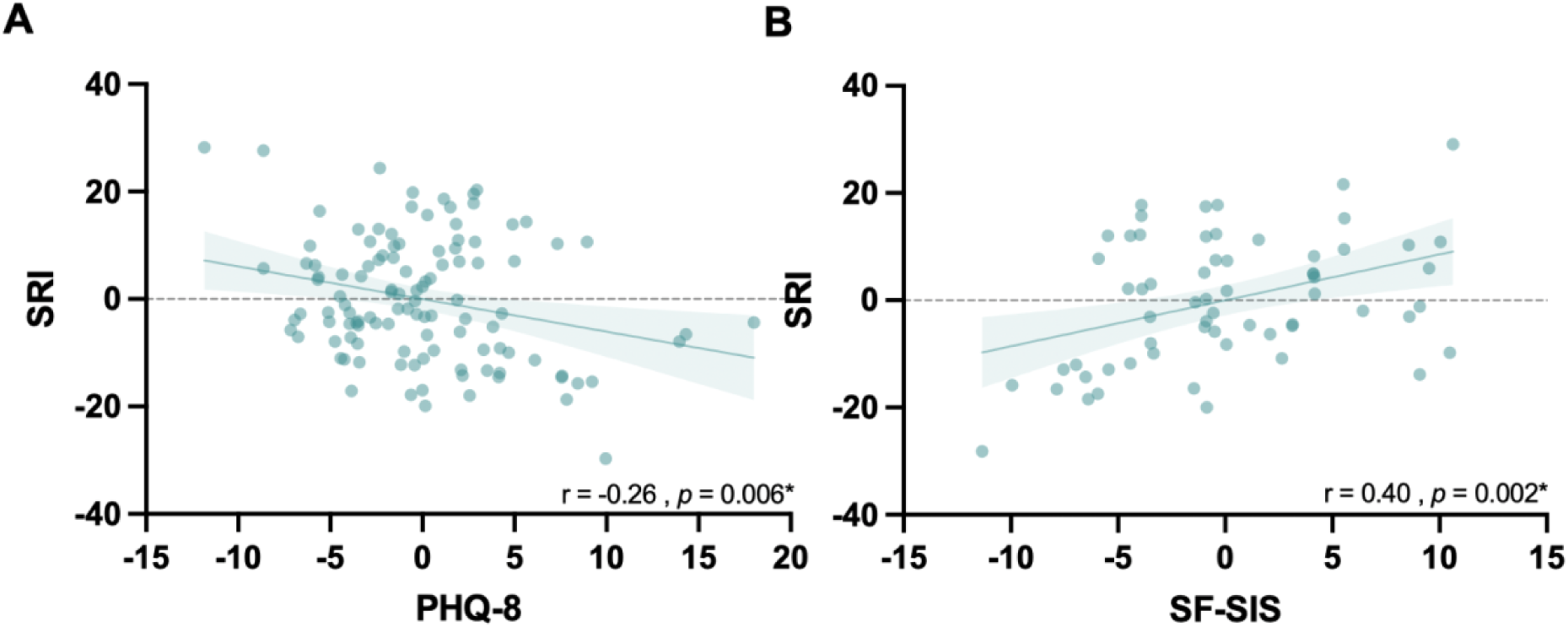
Plotted residuals controlled for age and sex to demonstrate partial correlations between **A.** SRI and the Patient Health Questionnaire depression scale (PHQ-8) (n=113 stroke survivors). **B.** SRI and the Stroke Impact Scale (n= 54 stroke survivors).

An exploratory objective was to investigate the correlation between SRI and quality of life (SF-SIS, n=54). There was a significant correlation (adjusted for age and sex; r = 0.40, *p* = 0.002), revealing that when stroke survivors have more regular sleep, their reported quality of life is higher (Figure 6B). However, there was no significant correlation between SRI and disability (mRS) or Time Since Stroke (supplementary S4).

## Discussion

The relationship between sleep and stroke is intricate, as sleep-wake disturbances are both a risk factor and potential outcome of stroke. In the present study we utilised a pre-existing actigraphy dataset from community-dwelling stroke survivors and established a significant disparity in sleep regularity between stroke survivors and healthy controls. We also found, for the stroke group alone, that more regular sleep was linked to longer overall nighttime sleep time, better self-reported sleep quality, less severe depression scores and better quality of life.

The group difference suggests that the stroke cohort exhibited greater irregularity in their sleep patterns. This result aligns with a growing body of literature suggesting that many aspects of sleep are negatively affected after stroke^2,28,29^ - which could result from several interrelated factors. Neurological damage resulting from a stroke can impair regions of the brain responsible for sleep regulation, leading to disruptions in the sleep-wake cycle. Psychological factors such as anxiety, depression, and stress, which are prevalent among stroke survivors, could also contribute to sleep irregularities^30^. These psychological factors have specifically been linked to post-stroke fatigue, a prevalent phenomenon that affects patients’ quality of life^31^. Post-stroke fatigue, as well as environmental factors and treatment schedules during hospitalisation and rehabilitation disrupt normal sleep routines, further complicating sleep regularity. Sleep irregularity has been a largely overlooked feature that may have clinical relevance. Specifically, the circadian system is crucial for maintaining overall health as it regulates numerous physiological functions, including the sleep-wake cycle^11^, which are instrumental in post-stroke recovery. Indeed, it has been shown that poor sleep can have a negative effect on rehabilitation outcomes^32^. Poor sleep following a stroke has further been associated with notable cognitive and attentional impairments, along with negative health and clinical outcomes^33^. However, when our analysis was repeated with subjective sleep quality (SCI) included in the model, the effect of group was no longer significant. We cannot rule out the influence of a sampling bias given that several of the included studies recruited participants with the intention of accessing a programme to try to improve their sleep problems. Therefore, we have also conducted a sensitivity analysis that can be found in supplementary materials (figure S5). However, given that stroke survivors typically experience worse self-reported sleep than controls^2^ this result may reflect inherent differences between these populations^3,34^, irrespective of recruitment techniques. The finding that better self-reported sleep is associated with more regular sleep patterns in stroke survivors is of particular interest as many studies report that subjective and objective measures of sleep do not necessarily align well^16,35^. One possible explanation for this association is that patients may be more attuned to irregularities in their sleep patterns than to other metrics, such as sleep efficiency. For example, disruptions to regular sleep schedules may manifest in noticeable behaviours like the need for daytime naps to compensate for inadequate or inconsistent nighttime rest. This heightened awareness of irregular sleep could account for the stronger subjective reporting of poorer sleep quality when sleep patterns are less regular. Furthermore, aspects of insomnia treatment (cognitive behavioural therapy for insomnia) aim to regulate sleep and wake patterns to improve patient perceptions of sleep quality. Future research should thus test whether improvements in sleep regularity can impact sleep quality, or vice versa, after stroke.

We also found that being of older age and male was associated with lower sleep regularity. This is consistent with prior research, with males generally having less regular sleep than females^36,37^. With regards to age, it has been proposed that changes in sleep with age are possibly caused by a diminished circadian control of sleep-wake cycles, and findings indicate that adjusting the circadian timing system could be a promising approach to mitigate declines in sleep quality and daytime alertness associated with aging^38^, which warrants further investigation. The SRI values observed in our study are lower than those reported in some larger population-based studies, which may be attributable to several factors. First, our study population is generally older than those used in many SRI studies, and as stated above, older adults may tend to exhibit lower SRI scores compared to younger, healthier individuals. Second, larger datasets, such as those from population cohorts like the UK Biobank, tend to capture a broader range of values, including more extreme scores, due to increased sample size and variability. Third, the specific type of actigraphy watch, smoothing algorithm and collection modality used (PIM in our study, versus ZCM in others) may also contribute to differences in SRI scores. While this difference in device sensitivity does not affect internal comparisons within our study, it should be considered when comparing our results to findings from studies using alternative actigraphy devices.

Within the stroke cohort only, a significant link was found between SRI and total sleep time (TST), where more regular sleep correlated with a longer actigraphy-derived total sleep time during the night. No associations were found within the healthy controls, but the strength of correlation did not differ significantly between the groups. Previous work has not found a correlation between TST and SRI in young healthy adults^13^ nor in a diverse sample of older adults^14^. Suboptimal sleep duration has been linked to an elevated risk of stroke occurrence^39^, and it remains to be seen whether sleep irregularity could also influence secondary stroke risk. We speculate that disruption to sleep after stroke could increase patients’ tendency to nap during the day, thereby diminishing sleep regularity.

Within the stroke cohort, the correlation between depression scores and SRI was significant (controlling for age and sex). This is consistent with previous studies that have revealed an association between sleep regularity and depression scores^14,40^. As the relationship between sleep and depression is complex and bi-directional^41^, it may be that irregular sleep patterns contribute to higher depression scores and/or that more severe depressive symptoms adversely affect sleep-wake patterns.

Finally, there was a significant relationship between sleep regularity and quality of life in stroke survivors, with higher sleep regularity indicating better quality of life. This finding aligns with previous work in other patient populations^42^, and is, to our knowledge, the first account reported in stroke. This is crucial, as earlier work has highlighted that sleep disorders can further reduce the quality of life in stroke patients, and diagnosing and treating sleep disturbance is vital to optimising functional outcomes and improving quality of life^43^. This finding presents an opportunity to investigate whether interventions targeting sleep regularity can enhance quality of life post-stroke.

We have demonstrated that the sleep regularity index could be a useful metric for post-stroke sleep research. When combined with other sleep metrics, SRI could potentially help address unresolved questions and provide new insights, as it uniquely accounts for sleep and wake patterns over the entire day and night. However, limitations of the present study should be mentioned. As the data were obtained from several studies investigating post-stroke sleep, each with distinct objectives, there was a potential sampling bias. All participants are community-dwelling stroke survivors, ranging from the sub-acute to the chronic phase (mostly in the chronic phase). Future research could attempt to replicate our findings, but in a less heterogeneous group and earlier post-stroke. It should further be noted that the sample consists mainly of stroke survivors who range from no to moderate disability, as measured by the mRS. There are very few participants with severe functional disability in the entire dataset. A general limitation in stroke research is that stroke survivors with very severe disabilities may not meet specific study criteria and may lack the physical or cognitive ability to participate, limiting generalisability across stroke. Finally, although we found no relationship between SRI and time since stroke, the study was cross-sectional. Future research should focus on examining changes in sleep regularity longitudinally, as this remains unexplored and the timescale by which sleep-wake problems occur post-stroke is unclear.

### Clinical Significance

To our knowledge, this study is the first to investigate the sleep regularity index in a stroke cohort. Stroke survivors exhibited greater irregularity in their sleep patterns than controls, and more irregular sleep was associated with shorter total sleep time, worse self-reported sleep and more severe depression. Most notably, there was evidence connecting sleep regularity with quality of life after stroke. Sleep regularity could be a novel target to improve post-stroke quality of life, but research is first needed to study changes in sleep regularity longitudinally, particularly to test for changes alongside recovery, and to identify potential methods for addressing sleep regularity problems.

## Acknowledgements

Tom Smejka and Ellie Macey who were involved in original data collection

## Author Contributions

**Conceptualization:** KBS, MKF

**Methodology:** KBS, MKF, AáVG, MW

**Data Collection:** MW, BR

**Data Analysis:** KBS, AáVG, MW

**Writing – Original Draft:** KBS

**Writing – Review & Editing:** KBS, AáVG, MW, BR, MKF, HJB

**Supervision:** MKF

**Funding Acquisition:** MKF, HJB

## Data availability

The code for all processing is open access and can be found on GitHub (https://github.com/KatSchruers/RISES). The data of this study are available from the corresponding author, MKF, upon reasonable request.

## Rights Retention

This research is funded in whole, or in part, by the Wellcome Trust [222446/Z/21/Z, 203139/Z/16/Z and 203139/A/16/Z]. For the purpose of open access, the author has applied a CCBY public copyright license to any Author Accepted Manuscript version arising from this submission.

