## Supplementary materials for "Sleep Regularity Index as a Novel Indicator of Sleep Disturbance in Stroke Survivors: A Secondary Data Analysis"

## S1

We utilised previously acquired actigraphy data from several studies conducted in our centre. In instances where a participant had taken part in multiple studies, the data with the longest recording or the most recent data was used (if duration was the same). Where participants had actigraphy at multiple timepoints in a single study, e.g. before and after an intervention, the baseline data were utilised.

**Table S1:** *Overview Data*

| Study | Participants | Inclusion criteria | Exclusion criteria | Measures |
|---|---|---|---|---|
| **Self-Reported and Objective Sleep Measures in Stroke Survivors with Incomplete Motor Recovery at the Chronic Stage[1]** | Stroke patients (69)<br>Healthy controls (70) | 1) Aged >18 years<br>2) Stroke >3 months prior<br>3) Self-reported difficulty using the upper limb<br>4) Able and willing to provide informed consent | 1) Neurological or psychological conditions other than stroke<br>2) Diagnosed sleep disorder prior to the stroke<br>3) Pre-stroke uncorrected visual impairment | Actigraphy<br>SCI<br>Sleep diary |
| **Improving sleep after stroke: a randomised controlled trial of digital cognitive behavioural therapy for insomnia[2]** | Stroke patients (84) | 1) Aged >18 years<br>2) > 3months post-stroke<br>3) Interested in improving their sleep<br>4) Living in the UK with reliable internet access<br>5) Able to understand verbal and written English<br>6) Able and willing to provide informed consent | 1) Serious clinical condition that could affect participation in the study, including scheduled surgery in the next 5 months<br>2) Currently undergoing a psychological treatment programme for insomnia<br>3) Pregnancy<br>4) Uncontrolled seizures<br>5) Untreated diagnosed obstructive sleep apnoea<br>6) Habitual shiftwork | Actigraphy<br>SCI<br>Sleep diary<br>PHQ-9<br>SF-SIS |
| **Improving sleep and learning in rehabilitation after stroke part 2[3]** | Stroke patients (24) | 1) Aged >18 years<br>2) Participant is willing and able to give informed consent for participation in the study 3) Clinical diagnosis of stroke affecting the upper limb, with | 1) Other neurological condition affecting movement (e.g. Parkinson's Disease, Multiple Sclerosis)<br>2) Diagnosed, untreated, sleep disorder (e.g. sleep apnoea) | Actigraphy<br>SCI<br>Sleep diary<br>PHQ-9<br>mRS |

| | | | | |
|---|---|---|---|---|
| | | sufficient movement to perform the motor learning task<br>4) Discharged from inpatient care<br>5) Interest in accessing a programme with the aim of improving sleep quality<br>6) Reliable access to the internet | 3) Uncontrolled seizures<br>4) Planned inpatient admission (e.g. for rehabilitation) in the next 4 months that would impact ability to engage with the Sleepio programme<br>4) Engagement in psychological therapy for insomnia in the past 12 months<br>5) Pregnant | |
| **Investigating consolidation of motor learning in the context of recovery after stroke[4]** | Stroke patients (15) | 1) Age > 18 years<br>2) Participant is willing and able to give informed consent for participation in the study OR a positive opinion from a consultee is provided by a family member or carer willing to provide personal consultee advice. | 1) Other neurological condition affecting movement (such as Parkinson's disease, Multiple Sclerosis) | Actigraphy<br>SCI<br>Sleep diary<br>PHQ-8<br>mRS |
| **The relationship between stroke lesion characteristics and sleep** | Stroke patients (19) | 1) Age > 18 years<br>2) Participant has a clinical diagnosis of stroke with an available structural brain scan obtained in a previous research study and has consented to be contacted about future studies.<br>3) Participant is willing and able to give informed consent for participation in the study.<br>4) Participant is discharged from inpatient hospital/rehabilitation care. | 1) The participant is a current shift-worker.<br>2) The participant has neurological or psychological conditions other than stroke with the researcher deems to preclude participation (e.g., Parkinson's Disease). | Actigraphy<br>SCI<br>Sleep diary<br>PHQ-9<br>mRS |

*Note.* Overview of all the studies that were be included in the current study, their inclusion and exclusion criteria, as well as all relevant measures for the current study. Abbreviations of measures include Patient Health Questionnaire (PHQ-8 and PHQ-9), Short form stroke impact scale (SF-SIS), Modified Rankin Scale (mRS), Sleep Condition Indicator (SCI). The PHQ-8 and PHQ-9 contain the same set up, however, PHQ-9 has one added question regarding suicidality, making it a nine-item depression scale. For the purpose of this study, the first 8 items are utilised only.

**S2**

*Actigraphy*

Actigraphy predicts sleep periods by assuming the body remains motionless during deep sleep, as opposed to wakefulness, using an activity count threshold of 20 (high sensitivity) in the MotionWare software (Camntech Ltd, UK). Actigraphy is considered to be a valid and reliable method for assessing sleep patterns in a home environment. Generally, actigraphy data can be collected through different modalities. The principal modalities include (1) Proportional Integrating Measure (PIM) mode captures high-resolution data by sampling the transducer signal at a rapid rate and calculating the area under the curve for each minute or 30-second interval. PIM serves as a measure of activity intensity or vigour and is employed in devices like the MotionWatch8 for comprehensive activity assessment. (2) Time Above Threshold (TAT) modality measures the movement duration within epochs which exceed the threshold. TAT provides a measure of overall time spent in motion. (3) Zero Crossing Method (ZCM), which quantifies motion frequency by counting the number of times per minute that a transducer signal (voltage) crosses a pre-set reference voltage threshold, typically set near zero. ZCM is the most commonly used modality, with a maximum motion frequency count of around 300. These modalities offer diverse approaches to capturing and analysing actigraphy data [5].

Actigraphy data were automatically classified as sleep or wake for each 30 second interval by the MotionWare programme. These data were used to calculate several sleep metrics, provided by the MotionWare programme. First, Total Sleep Time (TST), indicates the total duration spent in nightly sleep according to the epoch-by-epoch wake/sleep categorisation. Second, the sleep fragmentation index, which is calculated as the sum of the total time categorised as mobile, expressed as a percentage of the assumed sleep, and the number of immobile bouts which were ≤1 min in length, expressed as a percentage of the total immobile bouts. Third, the wake after sleep onset (WASO) refers to the number of minutes a participant was awake between sleep onset and sleep offset. Lastly, sleep efficiency is TST as a percentage of the overall time spent in bed. In other words, it is a measure that compares the amount of time spent sleeping to the total time generally spent in bed [1,5,6].

The Sleep Regularity Index (SRI) used extracted epoch data from actigraphy to calculate the percentage probability of an individual being in the same state (asleep vs. awake) at any two time-points 24 h apart, averaged across the study[7]. To guarantee a reliable calculation of the SRI, several criteria have been reported[8]. First, to guarantee sufficient data, participants must have at least five valid wear days (i.e. 5 full rounded days). Second, previous studies report that each participant should ideally have a weekend day included in the recording. Third, participants should not have > 2 hours of no recording (i.e. device off or not worn) in their duration of wearing the device[9].

*Smoothing*

Since actigraphy is an indirect method of measuring sleep, it may erroneously classify periods of inactivity during the day, such as when an individual remains motionless for short periods, as sleep. There are several algorithms reported in actigraphy literature that address this issue. Biegański, Stróż, Dovgialo, Duszyk-Bogorodzka and Durka [10] presented a unified framework untangling akin characteristics between the most common data scoring algorithms. It seems that most of the commonly used algorithms are equivalent to low-pass Finite Impulse Response (FIR) filters. This framework involves three main steps. Namely, summarizing continuous data into fixed time intervals (epochs), applying a mathematical operation called linear convolution using specific coefficients, and optionally adjusting the scoring for more accurate results. The first step involves downsampling, typically from 30 second epochs to one minute epochs. Additionally, the convolution coefficients used in various algorithms, applied to one minute epochs, effectively act as the low-pass filters that smooth the data. Among the most commonly used algorithms are Sadeh algorithm [11], Cole-Kripke algorithm [12] and University of California, San Diego (UCSD) algorithm [13]. The UCSD algorithm falls under the family of Cole-Kripke algorithms, that all act as low-pass FIR filters, but use different regression weights for separate actigraphy modalities [5,10]. The Sadeh algorithm was primarily validated in infants, children and adolescents [11] and has different characteristics to the Cole-Kripke family of algorithms, more details can be found in Biegański et al.[10] Overall, all actigraphy algorithms smooth the data, albeit in different ways, with the general goal of improving the accuracy of sleep/wake categorisation.

Algorithms that fall under the Cole-Kripke family are predominantly validated in adults [12-14], making them more appropriate for application to the participants of the present study. Nevertheless, multiple algorithms were tested to investigate the best fit (figures and further details below). The UCSD algorithm[5] was selected as the best fit for the current dataset. Since the current study has activity counts collected in PIM, whereas Cole-Kripke weights are generally based on ZCM.

The final algorithm used in the current study is shown in formula (2) with a scaling factor (*P*) of 0.10 to fit with the modality of the MotionWatch8. Consequently, $D < 1$ was scored as sleep, $D \geq 1$ was scored as wakefulness, and $A_{-4}$ to $A_{+2}$, were the activity scores for the 4 minutes preceding the minute of activity under consideration, the actual minute being scored, and the 2 succeeding minutes [13].

(2)

$$\mathrm{D} = \mathrm{P}(0.010\mathrm{A}_{-4} + 0.015\mathrm{A}_{-3} + 0.028\mathrm{A}_{-2} + 0.031\mathrm{A}_{-1} + 0.085\mathrm{A}_{0} + 0.015\mathrm{A}_{+1} + 0.010\mathrm{A}_{+2})$$

Lastly, to improve recording classification that includes both daytime and night-time activity (24h) Webster's rescoring rules were applied [14]. To summarise, Webster's rescoring rules indicate that (i) the first minute of a string of sleep scores should be rescored as wake score if preceded by 4 or more minutes of wake. That is, rescore 1 min of sleep to wake if the preceding 4 or more minutes were wake. (ii) 3 min of sleep should be rescored as wake if the preceding 10 min were wake. (iii) 4 min of sleep should be rescored as wake if the preceding 15 min were wake. (iv) if a period of 6 min or less that is scored as sleep and is surrounded (i.e. in both sides) by at least 15 min scored as wake, then rescore to wake (v) if a period of 10 min or less that is scored as sleep is surrounded (i.e. in both sides) by at least 20 min of wake, then rescore to wake [5,15]. Webster's rules were applied after the UCSD algorithm to reduce misclassification of sleep during the daytime period.

*Application of smoothing algorithm*

The original data was collected in 30-second epochs. These can be used to generate plots that represent the sleep/wake cycle and visualise the entire wear period, with lines that represent 30 seconds of sleep or wakefulness, in black and white respectively. See figure S1 for an example.

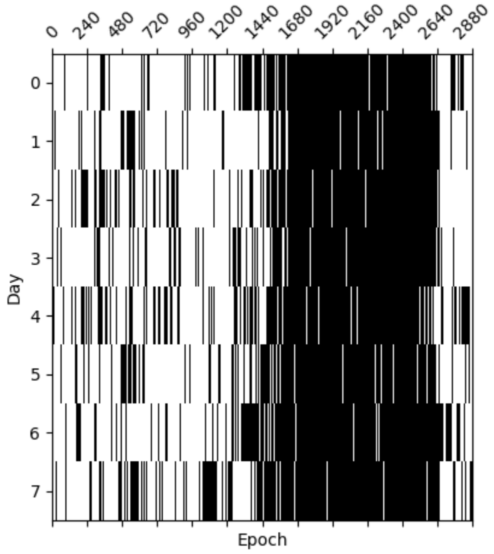


**Figure S1:** *Example of original raw data collected in 30-second epochs. The y-axis represents the 7 day wear period, while the x-axis represents a 24h shown in 30 second epochs. There are 2880 30 second epochs in a 24h period.*

However, since smoothing algorithms downsample to 1-minute epochs (collapsing of epochs), we applied this downsampling and tested multiple algorithms to identify the most suitable one for the current study's data. Our initial test used the Cole-Kripke algorithm. Unfortunately, this did not yield satisfactory results, an example can be seen in figure S2. This is likely due to the fact that our data is not collected in ZCM and does not fit well with the scaling factors used in the Cole-Kripke algorithm. It can be seen that this algorithm adds more wake in the general sleep period (indicated by disrupting white lines representing wake)

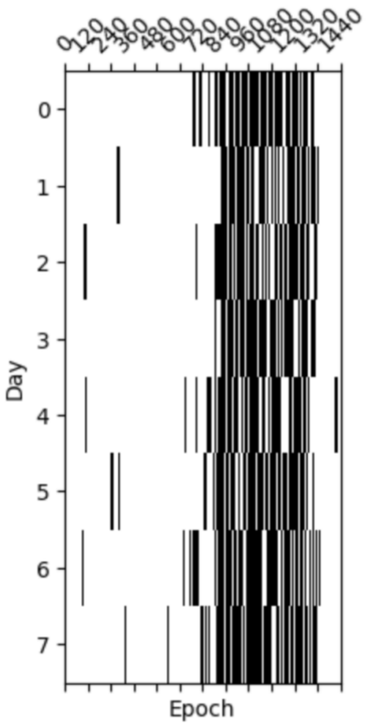


**Figure S2:** *Results from the Cole-Kripke algorithm. The y-axis represents the 7 day wear period, while the x-axis represents a 24h shown in 1 minute epochs. There are 1440 one minute epochs in a 24h period.*

We then tested the UCSD algorithm, which aligned better with our study's recording modality (PIM) but required a different scaling factor. This adjustment improved the results substantially and made the data more representative of what would be expected of a typical day-night cycle. An example can be seen in figure S3.

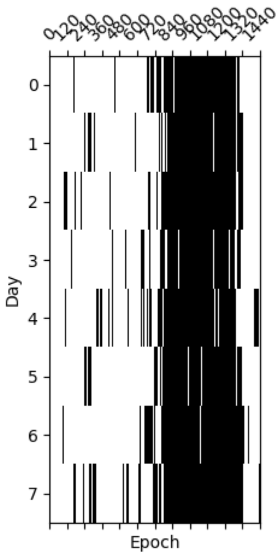

**Figure S3:** *Results from the UCSD algorithm with a scaling factor of 0.10. The y-axis represents the 7 day wear period, while the x-axis represents a 24h shown in 1 minute epochs.*

Finally, we applied Webster's rescoring rules to the data output from the UCSD algorithm, which also showed clear differences (see Figure S4). Throughout this process, we compared each stage against the raw data, and actigraph in MotionWare through visual inspection and selected the best option based on these comparisons.

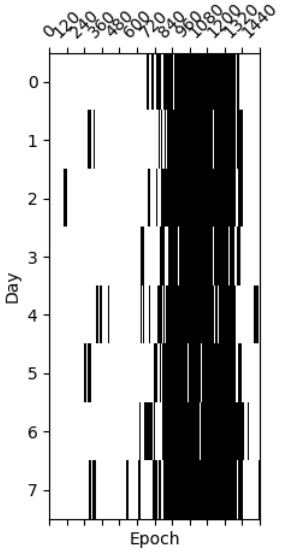


**Figure S4:** *Results after applying Webster's rules to the data smoothed by the UCSD algorithm. The y-axis represents the 7 day wear period, while the x-axis represents a 24h shown in 1 minute epochs.*

In conclusion, we felt that the most accurate representation of the data was achieved by utilizing the UCSD algorithm with an appropriately adjusted scaling factor, in conjunction with the application of Webster's rules. This approach effectively addressed inaccuracies in sleep stage detection during the day while avoiding excessive smoothing of the data.

The R code for all steps above is open access and can be found on GitHub (https://github.com/KatSchruers/RISES).

**S3**

*Statistical analyses*

For the linear model, all assumptions were checked, and regression diagnostics were inspected. These included the general linearity assumption, deviations from homoscedasticity, violations of normality and outliers. Residual plots indicated that the assumption of linearity was met. Additionally, partial residual plots for each predictor (Group, Age, and Sex) did not reveal any substantial deviations from linearity. To inspect deviations from homoscedasticity, residual plots were inspected, and a Levene's Test was conducted to assess the homogeneity of variances for the residuals in the linear model. The results indicated that the assumption of homogeneity of variances was met, $F(1,220)=0.66$, $p = 0.42$. Upon conducting a Shapiro-Wilk test on the residuals of the overall sample, which includes both the stroke and control groups (n=222), no significant violations of normality were detected (Shapiro-Wilk test: W = 0.9885, p-value = 0.0723). This suggests that the assumption of normality for residuals is upheld in the regression analysis.

The assumption of Pearson correlations were investigated both for the stroke group, and for the control group. For the stroke and control cohorts analysed separately, no variables showed non-linearity and inspection of plots revealed that the variables approximated a normal distribution, however, multiple variables violated the Shapiro wilk test of normality. Nevertheless, Havlicek and Peterson [16] showed that the Pearson correlations are robust against violations of normality. The authors concluded that the obtained distributions of person's correlation computed between scores from samples that violate normality (e.g., skewed) has little effect on the expected distribution especially with larger sample sizes (largest sample size within their study, n = 60). This was considered when examining all assumptions. Consequently, all correlations between variables were obtained using Pearson's correlations. Additionally, outliers were considered and upon inspection (z-score > 3), six outliers were detected for the stroke group across variables TST, WASO, fragmentation index and sleep efficiency and removed from the dataset for this analysis (thus, 157 stroke survivors were included). For the healthy controls, two outliers were detected and removed (58 healthy controls were included). For time since stroke, two outliers were detected and removed (final n = 160).

**S4**

**Results**

***Permutation testing***

Given the unequal sample sizes between stroke survivors (n=162) and the control group (n=60), we employed permutation testing to ensure the validity of the regression model results. Permutation testing is a non-parametric technique that evaluates the significance of an observed effect by repeatedly randomizing the data and recalculating the relevant test statistic. In this analysis, the stroke group was randomly subsampled to match the control group size and the group labels were shuffled across 10,000 iterations. For each iteration, the coefficient of the group variable was recalculated (stroke vs. control) from a linear regression model that controlled for age and sex. The observed coefficient for the group variable was −6.067, with a permutation-based $p$=0.0058. This indicates that the difference in SRI between stroke survivors and controls is statistically significant, even when accounting for the sample size disparity.

***Sensitivity analysis***

A sensitivity analysis was conducted to evaluate the robustness of the relationship between SRI scores and group (stroke vs. control), excluding two studies with potentially biased recruitment criteria (requiring participants to have a perceived sleep problem). In the revised model, the group effect remained in the expected direction, with stroke patients showing lower SRI scores than controls, although this effect did not reach statistical significance ($\beta$ = -3.64, 95% CI [-7.86, 0.57], t(130) = -1.71, $p$ = 0.090). The reduction in statistical power from the smaller sample likely contributed to the loss of significance. Age maintained a significant negative association with SRI ($\beta$ = -0.23, 95% CI [-0.37, -0.10], t(130) = -3.34, $p$ = 0.001), while sex was non-significant ($\beta$ = 3.89, 95% CI [-0.18, 7.97], t(130) = 1.89, p = 0.061). These results suggest that the directionality of the relationship remains consistent, indicating that stroke patients may still have a tendency towards lower SRI scores relative to controls. SRI values for each group are shown in figure S5.

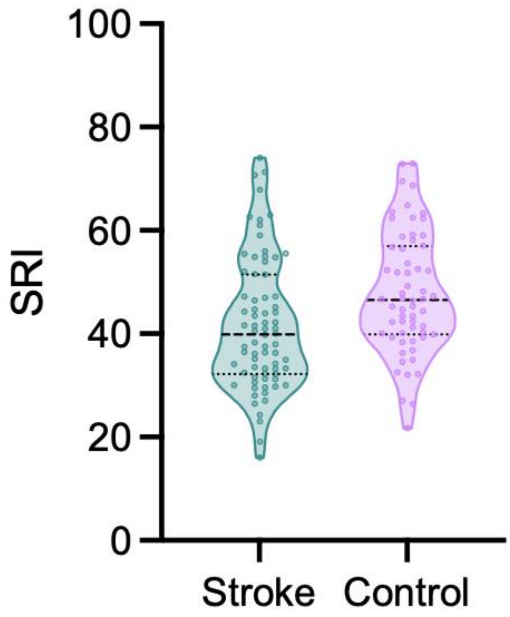


**Figure S5.** Violin plot showing the distribution of SRI scores in the stroke survivors and healthy controls for the subsample of stroke participants who were not recruited due to a perceived sleep problem. Higher SRI is indicative of more regular sleep. There was no significant effect of group, adjusting for age and sex ($p$ =0.090).

***Comparison strength of correlations between stroke cohort and healthy controls***

To determine if the strength of the raw correlations differed between groups, a Fisher's test was performed using the 'cocor.indep.groups' function of the cocor package in R. The test compared the correlation of SRI and TST between the stroke cohort and healthy controls, revealing Fisher's z = 0.878, $p$ = 0.380, indicating no difference in the strength of the correlation between the groups. Similarly, the strength of all other SRI-sleep metric correlations were tested between the groups and did not reveal any significant results (all $p > 0.05$).

**Exploratory analyses**

***SRI and mRS***

There was no significant correlation between SRI and mRS (r = 0.02, 95% CI [-0.25, 0.29], t(52) = 0.14, $p$ = 0.886), figure S6.

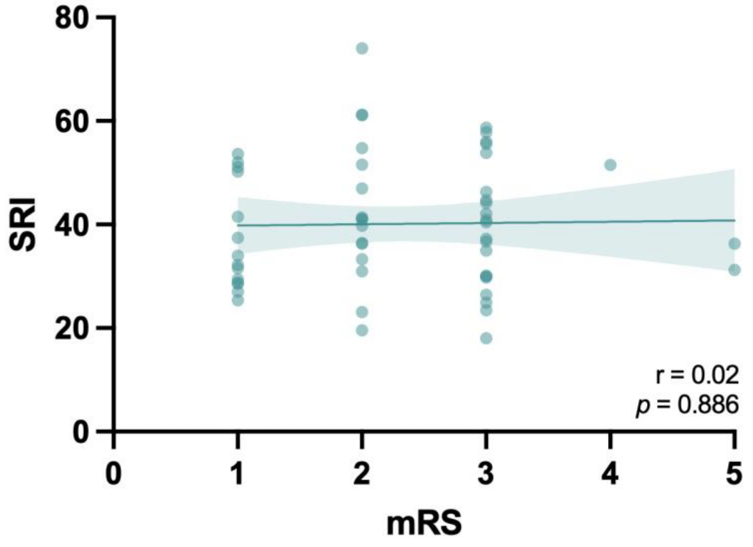


**Figure S6.** A scatterplot of the correlation between SRI and the modified Rankin Scale, available for a subset of 54 stroke survivors.

***SRI and Time Since Stroke***

There is no significant correlation between the SRI and Time since stroke (r = -0.04, 95% CI [-0.19, 0.12], t(158) = -0.46, $p$ = 0.646), figure S7.

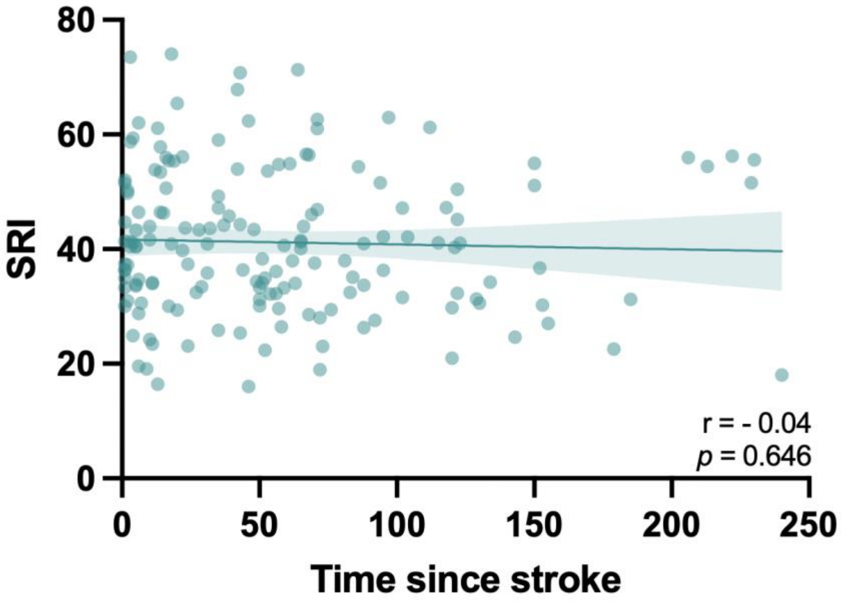


**Figure S7.** A scatterplot of the correlation between SRI and Time since stroke, 160 stroke survivors.
